# Multimorbidity Patterns and Associated Factors Among Middle-Aged and Older Adults in China: Evidence from the CHARLS Study

**DOI:** 10.64898/2026.03.31.26349821

**Authors:** Zijun Wang, Søren T. Skou, Yaolong Chen, Janne Estill

**Affiliations:** Center for Clinical Research and Evidence-Based Medicine, The First Hospital of Lanzhou University, 730000 Lanzhou, China; Research Unit of Evidence-Based Evaluation and Guidelines (2021RU017), Chinese Academy of Medical Sciences, School of Basic Medical Sciences, Lanzhou University, 730000 Lanzhou, China; Key Laboratory of Evidence Based Medicine of Gansu Province, 730000 Lanzhou, China; Institute of Health Data Science, Lanzhou University, 730000 Lanzhou, China; Evidence-based Medicine Center, School of Basic Medical Sciences, Lanzhou University, 730000 Lanzhou, China; The Research and Implementation Unit PROgrez, Department of Physiotherapy and Occupational Therapy, Central and West Zealand Hospital, 4200 Slagelse, Denmark; Department of Research, Central and West Zealand Hospital, 4200 Slagelse, Denmark; Center for Muscle and Joint Health, Department of Sports Science and Clinical Biomechanics, University of Southern Denmark, 5230 Odense, Denmark; Institute of Global Health, University of Geneva, 1202 Geneva, Switzerland

**Author notes:** Correspondence to: Prof. Yaolong Chen, Evidence-based Medicine Center, Lanzhou University, 199 Donggang West Rd, Chengguan District, 730000 Lanzhou, China. Email addresses Zijun Wang Søren T. Skou Yaolong Chen Janne Estill.

**Keywords:** Multimorbidity, latent class analysis, household survey

## Abstract

**Background:** Despite the growing global burden of multimorbidity, the patterns of disease combinations, have not been extensively categorized. We aimed to explore the predictors, health consequences, and patterns of discordant and concordant multimorbidity.

**Methods:** We used the 2018 China Health and Retirement Longitudinal Study (CHARLS), a representative database of adults aged ≥45 years from China. We conducted logistic regression analyses to assess the likelihood of having discordant (conditions from different disease systems) versus concordant (only cardiometabolic, or only respiratory diseases) multimorbidity, and to compare the health status and healthcare utilization between patients with discordant and concordant multimorbidity. Latent class analysis (LCA) was applied to both the entire sample and to patients with discordant multimorbidity to identify clusters of disease combinations.

**Results:** The sample included 1668 patients with concordant (mainly cardiometabolic), and 7306 patients with discordant, multimorbidity. Female patients, patients living in rural settings, former and current smokers, and patients engaging in high-intensity physical activity, were more likely to have discordant instead of concordant multimorbidity. Depression, limitations in daily activities, poor self-reported health, and frequent healthcare use were more common in patients with discordant than concordant multimorbidity. The LCA identified five clusters when all multimorbid patients were included (cardiometabolic, arthritis-digestive, respiratory, multisystem, and arthritis-hypertension classes), and four clusters when restricted to discordant multimorbidity (digestive, arthritis-cardiometabolic, respiratory, and multisystem classes).

**Conclusion:** Discordant multimorbidity is associated with poorer health and increased use of healthcare. Cardiometabolic diseases, arthritis, and digestive diseases have a central role in defining disease patterns.

## 1. Introduction

As the burden of multimorbidity increases globally, identifying patterns of disease combinations and implementing targeted management strategies is becoming increasingly important. In recent years, the number of epidemiological studies on multimorbidity—including prevalence, risk factors, and health outcomes—has grown rapidly.^1^ So far, there however exists no standardized method to classify multimorbidity. Compared with defining multimorbidity simply by the number of chronic conditions, an increasing number of studies have focused on the structural combinations of diseases and their potential relationships. This structural perspective helps reveal differences in clinical complexity across disease combinations and provides a basis for risk stratification and optimized management strategies. Some studies have categorized multimorbidity into concordant multimorbidity and discordant multimorbidity based on whether the coexisting diseases share similar pathophysiological mechanisms or management methods. This classification provides a clinically-relevant analytical framework for understanding potential synergies and conflicts of different disease combinations and offers ideas for integrating recommendations from multiple single-disease clinical practice guidelines in multimorbidity management.^2^

Concordant and discordant multimorbidity could provide a feasible foundation for a classification system for multimorbidity.^3^ However, both groups – particularly discordant multimorbidity – are themselves very heterogeneous, and the patterns of diseases need to be better understood. Moreover, most studies so far focus on populations determined by a single index disease, while systematic analyses based on the general population are scarce.^4,5^ Therefore, we aimed to explore the factors associated with discordant and concordant multimorbidity, assess the influence of discordance of the disease on the patients’ overall health, and identify common patterns of multimorbidity using clustering analysis of a large representative dataset of middle-aged and older adults from China.

## 2. Methods

### 2.1. Data source and sample

We obtained data from the 2018 China Health and Retirement Longitudinal Study (CHARLS), a household-based survey of Chinese individuals aged 45 years and older. The survey utilized a multistage probability sampling method to ensure national representativeness. The survey collected information on 14 chronic conditions: hypertension, dyslipidemia (high or low blood lipids), diabetes or elevated blood glucose, cancer, chronic lung disease (other than asthma), liver disease, heart disease, stroke, kidney disease, digestive diseases, memory-related diseases, psychiatric disorders, arthritis or rheumatism, and asthma. The national baseline survey was conducted in 2011, covering 150 county-level units and 450 village-level units, involving approximately 17,000 individuals from about 10,000 households. Follow-up surveys were conducted in 2013, 2015, 2018, and 2020.^6^

Due to the impact of the COVID-19 pandemic, the fourth wave (2020) of CHARLS contains substantial missing data for several key indicators. Therefore, this study used cross-sectional data from the third wave (2018), with 19,816 participants. Participants were included if they had complete information on key demographic and health variables, and had two or more of the 14 included chronic conditions. The dataset included demographic characteristics, lifestyle characteristics, health outcomes, and healthcare utilization (Table 1).

**Table 1.** Included variables and their categories.

| Variables | Categories |
| --- | --- |
| **Demographic characteristics** |  |
| Age | 45-54; 55-64; 65-74; $\geq 75$ years |
| Sex | Male; Female |
| Type of location | Urban; Rural |
| Marital status | Has spouse (including married and cohabiting); Does not have spouse (including separated, divorced, widowed, and unmarried) |
| Highest education level | No formal education; Primary school (including incomplete or home-based schooling given the participant is literate); Middle school; High school or above |
| **Lifestyle characteristics** |  |
| Smoking status | Nonsmoker; Current smoker; Former smoker |
| Alcohol consumption | Drinking any alcohol in the past year; Not drinking any alcohol in the past year (based on participants' reports of consuming alcoholic beverages) |
| Physical activity | Engaging in at least 10 minutes of exercise per week of varying intensity, categorized as: High intensity activities; Moderate-intensity activities; Low-intensity activities; No activity. |
| **Health outcomes** |  |
| Depression status | Assessed using the 10-item Center for Epidemiologic Studies Depression Scale (CESD-10). The total score ranges from 0 to 30, with higher scores indicating more severe depressive symptoms. Participants were categorized as either experiencing depression (CESD-10 $\geq 12$ points) or not experiencing depression (CESD-10 $< 12$ points).<sup>[7]</sup> |
| Activity of Daily Living (ADL) | Assessed using six items: bathing, dressing, eating, getting in/out of bed, using the toilet, and controlling urination. The assessment categorized participants as either having no ADL limitations (experiencing no difficulty in any of the above activities) or having ADL limitations (experiencing difficulty in any one of the above activities).<sup>[8]</sup> |
| Self-reported health status | Very good; Good; Fair; Poor; Very poor |
| **Healthcare utilization** |  |
| Doctor visit/outpatient in the past month | Yes; No |
| Hospital stay in the past year | Yes; No |
| Satisfaction with local health care services | Not at all satisfied; Not very satisfied; Somewhat satisfied; Satisfied; Completely satisfied |

### 2.2. Definition of concordant and discordant multimorbidity

In this study, concordant multimorbidity was defined as either the coexistence of two or more cardiometabolic diseases (hypertension, diabetes, dyslipidemia, heart disease, stroke, liver disease, or kidney disease), or the coexistence of asthma and another chronic lung disease, without other conditions. Any other combination of diseases was defined as discordant multimorbidity.

### 2.3. Statistical analysis

Weighted data with the original survey weights were used for all analyses. We first explored the associations between different variables and the type of multimorbidity (concordant vs. discordant). Descriptive data were presented as frequencies (unweighted counts) and weighted percentages. Chi-square tests were used to compare differences in demographic and lifestyle characteristics between the two groups. Subsequently, all available demographic variables and lifestyle factors were entered into multivariable logistic regression models to adjust for potential confounding and identify factors independently associated with the type of multimorbidity. P ≤0.05 was considered statistically significant.

To examine the potential effects of concordant vs. discordant multimorbidity patterns on health outcomes and healthcare utilization, logistic regression analyses (binomial or ordinal, depending on the type of the outcome variable) were conducted for six outcomes: depression status, limitations in activities of daily life (ADL), self-reported health status, doctor or outpatient visit in past month, hospitalization in past year, and satistfaction in local health services. Three models were constructed: univariable model (Model 1); multivariable model adjusted for age, sex, marital status, and type (urban/rural) of location (Model 2); and multivariable model further adjusted for smoking status, alcohol consumption, and physical activity in addition to the variables included in Model 2 (Model 3).

Next, we used latent class analysis (LCA) to identify multimorbidity patterns across the 14 chronic conditions. LCA is a model-based clustering method suitable for multivariate categorical data and is commonly used to identify latent subgroups within populations. Models with 2 to 8 classes were fitted and compared using Akaike Information Criterion (AIC), Bayesian Information Criterion (BIC), log-likelihood, and entropy. The optimal number of classes was selected based on model fit, parsimony, and interpretability. We first considered BIC in determining the optimal cluster, and compared it to the other indicators. If the results of these indicators differed, or if there were very small classes (<5%), we considered also the three other indicators, discussed the results among the authors, and decided the optimal number by consensus.^7^ Two separate LCAs were conducted: (1) including all individuals with two or more chronic diseases to identify overall multimorbidity patterns; (2) among individuals excluding concordant multimorbidity, to identify patterns of discordant multimorbidity.

The LCA was performed in R (version 4.5.0). All other analyses and data management were conducted using STATA software (version 15.1).

## 3. Results

### 3.1. Baseline characteristics

After excluding patients with missing key variables or only one condition, 8974 individuals were included in the analysis (Figure 1). The characteristics of participants are shown in Table 2. The median age of the study population was 63 (interquartile range [IQR] 55-69) years. Slightly more than half (52.5%, n=4889) of the participants were women, and most participants lived in rural areas and urban areas were balanced in proportion (n = 5,465, 50.1% vs. n = 3,509, 49.9%). Regarding lifestyle behaviors, 57.4% were non-smokers (n=5165), and 66.7% did not consume any alcohol during the past year (n=6202). About nine our of ten participants engaged regularly in at least some level of physical activity; 2716 (26.6%) engaged in high-intensity activities. Regarding health outcomes, 28.2% had depressive symptoms (n=2768), 22.9% had limitations in activities of daily living (n=2225), and 35.4% reported poor or very poor health (n=3365). Around 20% of the participants had used outpatient or home healthcare services in the past month, or had been hospitalized in the past year. The most prevalent chronic conditions were arthritis (n=5304, 55.6%), hypertension (n=5169, 57.9%) and digestive diseases (n=4224, 45.0%). The clear majority of the patients (n=7306; 81.4%) had discordant multimorbidity, 1610 (17.9%) had concordant cardiometabolic multimorbidity, and 58 (0.6%) concordant respiratory multimorbidity. There were statistically significant differences in the distribution of sex, type of location, marital status, educational level, alcohol consumption, and physical activity between the patients with concordant and discordant multimorbidity (Table 2).

**Table 2.**
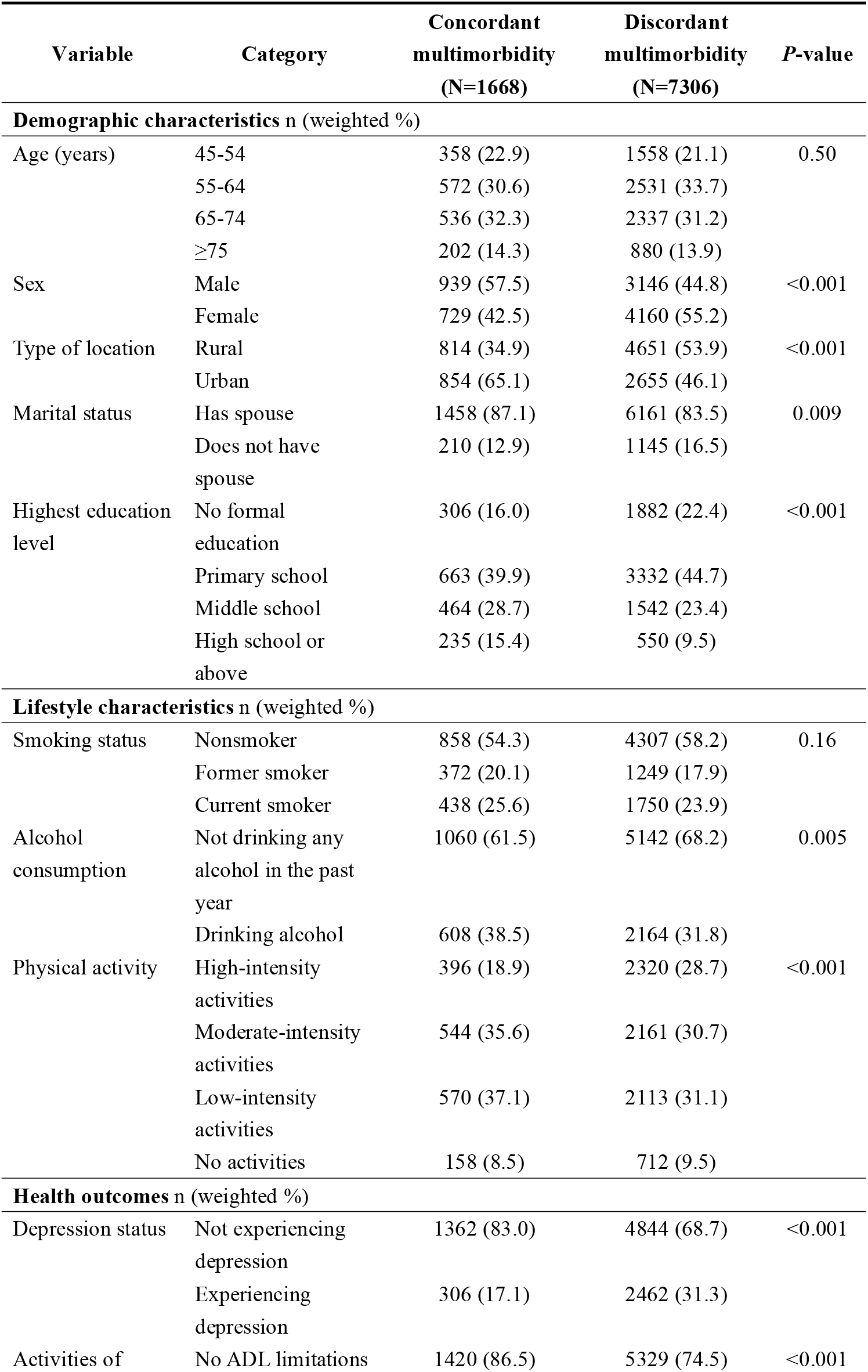
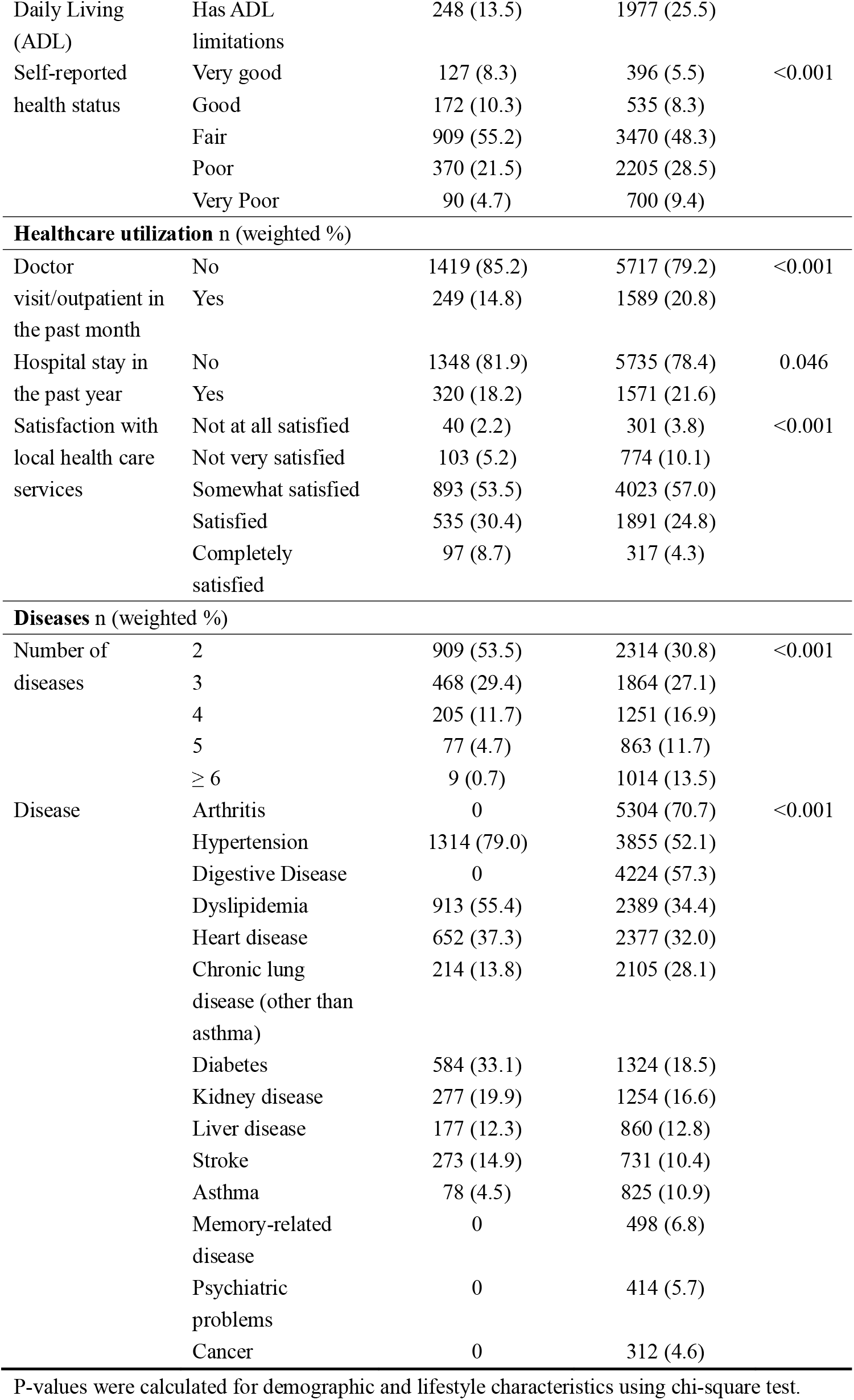
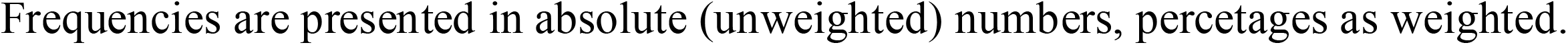
Baseline characteristics of the included patients with concordant and discordant multimorbidity.

**Figure 1.**
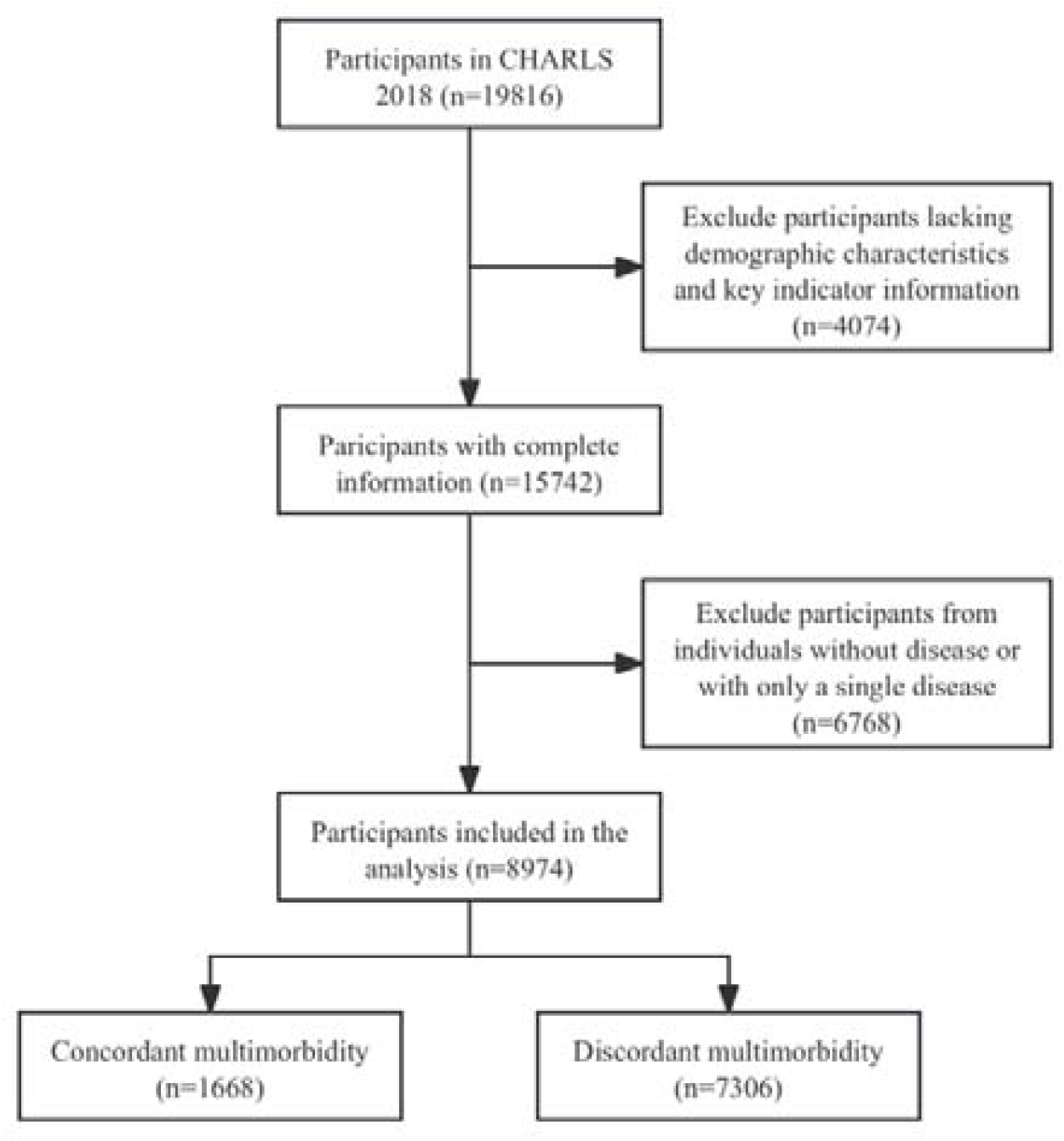
Flowchart of the participant inclusion

### 3.2. Predictors of discordant multimorbidity

According to the results of the multivariable regression analysis, women were significantly more likely than men to have discordant instead of concordant multimorbidity (odds ratio [OR] = 2.17, 95% confidence interval [CI]: 1.58–2.98, *P* < 0.001; Table 3). Participants living in rural areas were also more likely to have discordant multimorbidity compared with those living in urban areas (OR = 1.89, 95% CI: 1.58–2.25, *P* < 0.001). Compared with nonsmokers, both former (OR = 1.48, 95% CI: 1.07–2.04) and current (OR = 1.47, 95% CI: 1.04–2.06) smokers showed an elevated likelihood of discordant multimorbidity (*P* = 0.048). Compared with individuals engaging in high-intensity physical activity, those participating in moderate-intensity activity, low-intensity activity, or no physical activity were significantly less likely to have discordant multimorbidity (OR = 0.62, 95% CI: 0.49–0.79; OR = 0.64, 95% CI: 0.49–0.82; and OR = 0.70, 95% CI: 0.53–0.93, respectively; *P* = 0.001). In contrast, age, marital status, educational level, and alcohol consumption were not significantly associated with multimorbidity type in the multivariable analysis.

**Table 3.** Multivariable logistic regression analysis of factors associated with having discordant vs. concordant multimorbidity.

| Variables | Discordant vs. concordant multimorbidity |  |  |
| --- | --- | --- | --- |
|  | OR | 95%CI | P-value |
| **Age (years)** |  |  |  |
| 45-54 | 1 (ref) |  | 0.255 |
| 55-64 | 1.26 | 0.96-1.65 |  |
| 65-74 | 1.07 | 0.80-1.45 |  |
| ≥75 | 1.14 | 0.81-1.60 |  |
| **Sex** |  |  |  |
| Male | 1 (ref) |  |  |
| Female | 2.17 | 1.58-2.98 | <0.001 |
| **Type of location** |  |  |  |
| Urban | 1 (ref) |  |  |
| Rural | 1.89 | 1.58-2.25 | <0.001 |
| **Marital status** |  |  |  |
| Has spouse | 1 (ref) |  |  |
| Does not have spouse | 1.18 | 0.95-1.46 | 0.124 |
| **Highest education level** |  |  |  |
| No formal education | 1 (ref) |  | 0.092 |
| Primary school | 1.11 | 0.87-1.42 |  |
| Middle school | 0.90 | 0.67-1.21 |  |
| High school or above | 0.77 | 0.54-1.09 |  |
| **Smoking status** |  |  |  |
| Nonsmoker | 1 (ref) |  | 0.048 |
| Former smoker | 1.48 | 1.07-2.04 |  |
| Current smoker | 1.47 | 1.04-2.06 |  |
| **Alcohol consumption** |  |  |  |
| Not drinking any alcohol in the past year | 1 (ref) |  |  |
| Drinking alcohol | 0.98 | 0.80-1.19 | 0.802 |
| **Physical activity** |  |  |  |
| High-intensity activities | 1 (ref) |  | 0.001 |
| Moderate-intensity activities | 0.62 | 0.49-0.79 |  |
| Low-intensity activities | 0.64 | 0.49-0.82 |  |
| No activities | 0.70 | 0.53-0.93 |  |

Odds Ratio, OR; 95% Confidence Interval, 95%CI
P values calculated by adjusted Wald test

### 3.3. Effects of discordant multimorbidity on health and healthcare utilization

Stratified logistic regression analyses were performed to examine the associations between the five multimorbidity patterns and health outcomes as well as healthcare utilization, using models constructed based on demographic and lifestyle characteristics. The results are presented in Table 4.

**Table 4.** Logistic regression analysis of the effects of concordant and discordant multimorbidity on health outcomes and healthcare utilization.

| Variables | Model 1 (univariable) |  |  | Model 2 (multivariable)* |  |  | Model 3 (multivariable)** |  |  |
| --- | --- | --- | --- | --- | --- | --- | --- | --- | --- |
|  | OR | 95%CI | P-value | OR | 95%CI | P-value | OR | 95%CI | P-value |
| **Health-related Outcomes** |  |  |  |  |  |  |  |  |  |
| Depression status: yes vs. no |  |  |  |  |  |  |  |  |  |
| Concordant multimorbidity | 1 (ref) |  |  | 1 (ref) |  |  | 1 (ref) |  |  |
| Discordant multimorbidity | 2.22 | 1.73-2.84 | <0.001 | 1.85 | 1.45-2.36 | <0.001 | 1.82 | 1.42-2.33 | <0.001 |
| Activity of Daily Living (ADL): limitations vs. no limitations |  |  |  |  |  |  |  |  |  |
| Concordant multimorbidity | 1 (ref) |  |  | 1 (ref) |  |  | 1 (ref) |  |  |
| Discordant multimorbidity | 2.20 | 1.79-2.70 | <0.001 | 1.99 | 1.62-2.43 | <0.001 | 2.03 | 1.66-2.50 | <0.001 |
| Self-reported health status: 5-level scale (OR from ordinal logistic regression, poorer vs. better health) |  |  |  |  |  |  |  |  |  |
| Concordant multimorbidity | 1 (ref) |  |  | 1 (ref) |  |  | 1 (ref) |  |  |
| Discordant multimorbidity | 1.62 | 1.37-1.93 | <0.001 | 1.48 | 1.24-1.76 | <0.001 | 1.50 | 1.27-1.79 | <0.001 |
| **Healthcare Utilization** |  |  |  |  |  |  |  |  |  |
| Doctor visit/outpatient in the past month: yes vs. no |  |  |  |  |  |  |  |  |  |
| Concordant multimorbidity | 1 (ref) |  |  | 1 (ref) |  |  | 1 (ref) |  |  |
| Discordant multimorbidity | 1.51 | 1.25-1.84 | <0.001 | 1.47 | 1.21-1.78 | <0.001 | 1.47 | 1.21-1.78 | <0.001 |
| Hospital stay in the past year: yes vs. no |  |  |  |  |  |  |  |  |  |
| Concordant multimorbidity | 1 (ref) |  |  | 1 (ref) |  |  | 1 (ref) |  |  |
| Discordant multimorbidity | 1.24 | 1.00-1.53 | 0.046 | 1.28 | 1.03-1.59 | 0.024 | 1.29 | 1.04-1.61 | 0.022 |
| Satisfaction with local health care services: 5-level scale (OR from ordinal logistic regression, more vs. less satisfied) |  |  |  |  |  |  |  |  |  |
| Concordant multimorbidity | 1 (ref) |  |  | 1 (ref) |  |  | 1 (ref) |  |  |
| Discordant multimorbidity | 0.59 | 0.48-0.73 | <0.001 | 0.60 | 0.49-0.73 | <0.001 | 0.61 | 0.51-0.74 | <0.001 |

Odds Ratio, OR; 95% Confidence Interval, 95%CI
\*Model 2: adjusted for age, sex, type of location, and marital status.
\*\*Model 3: adjusted for age, sex, type of location, marital status, smoking, alcohol consumption, and physical activity.

Having discordant multimorbidity (as opposed to concordant) was significantly associated with depressive symptoms (OR = 1.82, 95% CI: 1.42–2.33, *P* < 0.001 in Model 3), limitations in ADL (OR = 2.03, 95% CI: 1.66–2.50, *P* < 0.001 in Model 3), and poorer self-rated health (OR = 1.50, 95% CI: 1.27–1.79, *P* < 0.001 in Model 3). The associations were robust and statistically significant across all three models.

Discordant multimorbidity was also significantly associated with higher utilization of outpatient (OR = 1.47, 95% CI: 1.21–1.78, *P* < 0.001 in Model 3) and home-based services, increased hospitalization (OR = 1.29, 95% CI: 1.04–1.61, *P* < 0.001 in Model 3), and lower satisfaction with healthcare services (OR = 0.61, 95% CI: 0.51–0.74, *P* < 0.001 in Model 3). The significance, magnitude and direction of the associations did not differ across the three models.

### 3.4. Latent class analysis of multimorbidity patterns

In the first LCA including all participants with multimorbidity (concordant or discordant), the seven-class model yielded the lowest BIC. However, the five-class model was finally selected as the optimal one because it provided parsimonious and clinically interpretable multimorbidity patterns, had no class with very small sample sizes (<5%), and maintained similar entropy compared with models with a larger number of classes. Detailed model fit indices are presented in Table A.1 in the Appendix. Based on the proportional distribution and prevalence patterns of chronic diseases within each latent class, descriptive labels were assigned to summarize the main disease combinations in each pattern to facilitate interpretation and discussion of the results (Figure 2).

**Figure 2.**
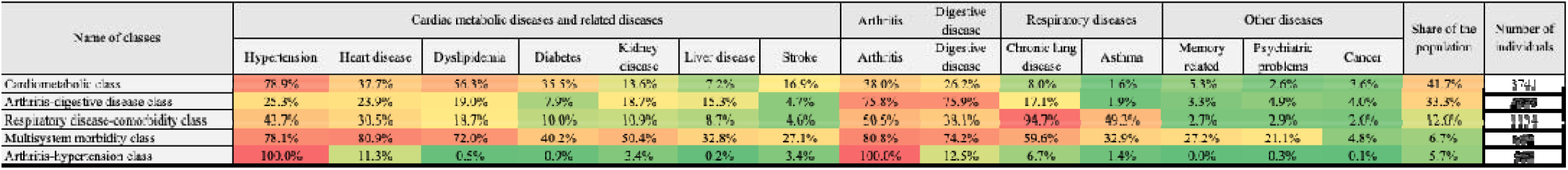
Results of the latent class analysis of patients with multimorbidity

Pattern 1 (n=3741, 41.7%) was characterized primarily by cardiometabolic diseases, including hypertension (78.9%), dyslipidemia (56.3%), heart disease (37.7%), and diabetes (35.5%). This pattern was labeled “Cardiometabolic class” and represented the most prevalent pattern. Pattern 2 (n=2989, 33.3%) was characterized by a high prevalence of arthritis (75.8%) and digestive diseases (75.9%), and was labeled “Arthritis-digestive disease class”. Pattern 3 (n=1134, 12.6%) was characterized by arthritis (50.5%) and respiratory diseases, including chronic lung disease (94.7%) and asthma (49.3%). This pattern was labeled “Respiratory disease-comorbidity class”. Pattern 4 (n=602, 6.7%) showed high prevalence across multiple disease systems, including cardiometabolic diseases (hypertension 78.1%, heart disease 80.9%, dyslipidemia 72.0%, and kidney disease 50.4%), arthritis (80.8%), digestive diseases (74.2%), and respiratory diseases (chronic lung disease 59.6% and asthma 32.9%). This pattern was labeled “Multisystem morbidity class”. Finally, pattern 5 (n=508, 5.7%) was characterized by the coexistence of hypertension (100.0%) and arthritis (100.0%), and was labeled “Arthritis-hypertension class”.

For participants with discordant multimorbidity (N=7306), the four-class model was selected as the best fit, for the same reasons as the five-class model in the first analysis. Detailed model fit statistics are shown in Table A.2 in the Appendix.

Many similar patterns as in the first analysis were observed also in the second one (Figure 3). The four clusters were labeled as “Digestive disease-comorbidity class” (n=2753, 37.7%), “Arthritis-cardiometabolic class” (n=1959, 26.8%), “Respiratory disease-comorbidity class” (n=1572, 21.5%), and “Multisystem morbidity class” (1022, 14.0%).

**Figure 3.**
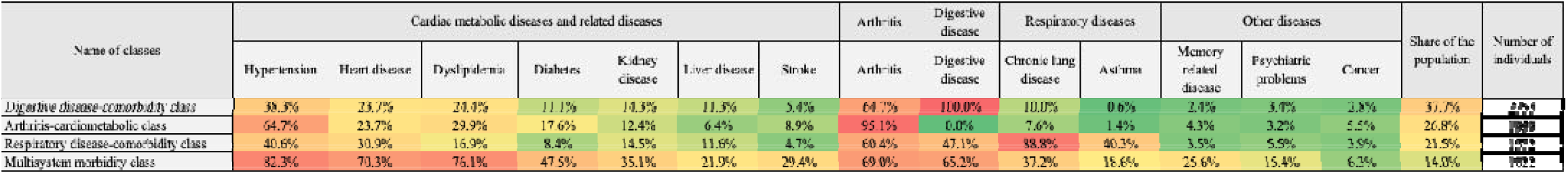
Results of the latent class analysis of patients with discordant multimorbidity

## 4. Discussion

### 4.1. Summary of main findings

This study used cross-sectional data from the third wave of the CHARLS study to examine disease combination patterns among middle-aged and older adults with multimorbidity in China. We compared the characteristics of individuals with concordant and discordant multimorbidity, and explored their associations with health outcomes and healthcare utilization. With the help of two LCA analyses, we identified five multimorbidity classes. Four of the classes were centered around cardiometabolic diseases, arthritis, digestive diseases, and respiratory diseases, and one class contained patients with multiple diseases from different disease systems. Our results further demonstrate that discordant multimorbidity accounted for the clear majority of the multimorbid population, and almost half of the overall population aged 45 and above. Discordant multimorbidity was significantly more common among women, rural residents, and smokers, and individuals engaging in high-intensity physical activity than their counterparts. Individuals with discordant multimorbidity were more likely to experience depressive symptoms, limitations in activities of daily living, and poorer self-rated health, as well as higher rates of outpatient visits and hospitalizations, and lower satisfaction with healthcare services, compared with those with concordant multimorbidity.

### 4.2. Comparison with previous studies

LCA is a commonly used method for identifying multimorbidity patterns. A 2024 study by Li Xu and colleagues used LCA based on the 2011 and 2015 CHARLS datasets and identified five multimorbidity classes among middle-aged and older adults in China: cardiometabolic diseases, cardiometabolic diseases with musculoskeletal diseases, musculoskeletal diseases, respiratory diseases, and non-specific multimorbidity.^8^ Similarly, a 2023 study by Yaqin Zhong et al. using the 2018 CHARLS data identified five disease patterns: relatively healthy class, vascular class, respiratory class, stomach-arthritis class, and multisystem morbidity class.^9^ Overall, the multimorbidity patterns identified in the present study are broadly consistent with those reported in previous research, particularly regarding the clustering of cardiometabolic diseases, suggesting that these disease combinations exhibit stability and persistence within the population. However, methodological differences exist among studies. Xu et al. incorporated objective indicators such as body mass index in addition to self-reported diseases.^8^ Zhong et al. highlighted the clustering characteristics of vascular diseases, emphasizing the coexistence of hypertension, dyslipidemia, diabetes, and cardiovascular diseases. Nevertheless, differences in study population definitions and sample composition should be noted.^9^ Xu et al. included only individuals aged 60 years or older with available physical examination data, which may underestimate multimorbidity patterns among middle-aged adults.^8^ Zhong et al. did not exclude individuals with only a single disease in the LCA analysis, which may affect the identification of “real multimorbidity patterns”.^9^ In contrast, the present study included all individuals aged 45 years and older with multimorbidity in the 2018 CHARLS dataset and explicitly excluded individuals with only a single chronic condition, thereby focusing more specifically on the structure of disease coexistence. Moreover, we considered also an alternative approach where individuals with concordant multimorbidity were first excluded. The patterns identified in the LCA were however similar: in both situations, all but one classes were centered around certain conditions – particularly arthritis, cardiometabolic diseases, and digestive disease – while one cluster in both analyses was formed by those with complex multisystem morbidity, These findings further confirm the central role of cardiometabolic diseases in the multimorbidity structure among middle-aged and older adults in China.

Although the number of studies is limited, differences between concordant and discordant multimorbidity and their effects on health outcomes have received increasing attention. A study from South Africa reported that 72% of multimorbidity cases were concordant and 28% discordant.^10^ A study conducted in India on diabetes and its comorbidities reached similar conclusions, showing that concordant multimorbidity was more prevalent than discordant multimorbidity.^11^ In contrast, in the present study, individuals classified as having concordant multimorbidity represented only one-quarter of those with discordant multimorbidity. This discrepancy may be explained by differences in study populations and disease inclusion criteria. The present study analyzed combinations of 14 self-reported chronic non-communicable diseases in the CHARLS dataset, whereas the above studies also included infectious diseases such as HIV and tuberculosis, reflecting regional disease characteristics. Moreover, our study included everyone with at least two conditions, therefore taking the patient-centered “multimorbidity” perspective instead of the “comorbidity” perspective focusing on a certain index disease like diabetes. The concept of concordant and discordant multimorbidity was indeed originally developed in the context of diabetes and its comorbidities^3^, and most existing studies continue to focus on populations with diabetes. From the perspective of diabetes-related comorbidities, conditions sharing similar pathophysiological mechanisms with diabetes are naturally more common. However, as demonstrated by our results, while the cardiometabolic cluster (including also diabetes amongst other diseases) takes a major share of the overall multimorbidity burden, these conditions also co-occur often with other diseases, calling for more focus on discordant and complex combinations.

Regarding health outcomes and healthcare utilization, individuals with discordant multimorbidity experienced poorer health outcomes and higher healthcare utilization, which is consistent with most previous research. A previous study by Ricci-Cabello et al. suggested that as individuals with concordant multimorbidity benefit from synergies between the clinical management approaches, the quality of care is likely to be higher than among those with discordant multimorbidity.^12^ A study by Lin et al. classified diabetes-related multimorbidity into five categories: diabetes with concordant comorbidities only, with discordant comorbidities only, with both concordant and discordant comorbidities, with any dominant disease, and diabetes alone.^5^ Analysis of mean healthcare expenditures showed that diabetes with any severe, clinically dominant comorbidity had the highest costs (USD 38,168), which was 87% higher than those for individuals with both concordant and discordant conditions (USD 20,401), and over fourfold higher than those with discordant conditions only (USD 9,173) or concordant conditions only (USD 9,000). All these costs were significantly higher than those for patients with diabetes alone (USD 3,365). Taken together, these findings indicate that although multiple factors influence health outcomes and healthcare utilization among patients with multimorbidity, differences between concordant and discordant multimorbidity are substantial and consistently observed across studies.

### 4.3. Implications for practice

Distinguishing between concordant and discordant multimorbidity has important implications for optimizing clinical decision-making and patient management. For example, in the case of concordant multimorbidity involving cardiometabolic diseases, although complex disease combinations may pose challenges for patients and primary care providers, experienced cardiovascular specialists can often manage such patients relatively effectively due to the shared or related pathophysiological mechanisms among these diseases.^13^ Moreover, common concordant disease combinations are often addressed in single-disease clinical practice guidelines, which can also streamline the care. In practice, healthcare providers should place greater emphasis on early prevention and control of disease progression in patients with concordant multimorbidity to reduce the occurrence of additional or more severe conditions.^14^ In contrast, diseases involved in discordant multimorbidity often lack direct or well-established associations, and the treatment approaches of the different comorbidities may differ or even conflict with each other. As a result, their management is more complex and requires broader professional knowledge and additional healthcare resources. For patients with relatively mild discordant multimorbidity, primary care physicians may be able to provide initial management. However, as disease severity increases, a single specialist may be unable to provide comprehensive care, and patients may need to consult multiple specialists, increasing the burden on both individuals and the healthcare system. Therefore, in response to increasingly complex healthcare needs, healthcare systems should shift from the traditional single-disease, specialty-oriented care model to a patient-centered and integrated care model. For patients with discordant multimorbidity, management should not simply involve the accumulation of treatments from multiple specialties; rather, it should rely on multidisciplinary collaboration, comprehensive assessment, and integrated management, emphasizing shared decision-making between clinicians and patients and providing individualized, coordinated, and sustainable care.^15^

### 4.4. Limitations

This study has several limitations. First, the analysis was based on CHARLS data, which is representative of the population of China aged 45 year or above, but the results may not be generalizable to other countries or younger age groups. Second, the 14 chronic diseases included in this study were based on self-reported information, which may be subject to recall bias. Third, there is still no unified definition for concordant and discordant multimorbidity. The classification we used relied primarily on previous literature and own judgment. For example, the cardiometabolic cluster was formed by combining cardiovascular diseases with conditions from other systems such as diabetes and liver and kidney disease. Finally, the 14 included diseases, despite being generally common, constitute only a small part of the existing chronic conditions. Therefore, analyses with a broader geographic focus, inclusion of more diseases, and a large sample size are still needed to understand better the patterns and mechanisms behind multimorbidity.

## 5. Conclusion

In this analysis, we used latent class analysis to cluster the patterns of multimorbidity using the 2018 wave of the CHARLS database, and furthermore systematically compared differences in demographic characteristics, health outcomes, and healthcare utilization between individuals with concordant and discordant multimorbidity. The results indicate that cardiometabolic multimorbidity and the combination of arthritis and digestive diseases represent the most common disease patterns for concordant and discordant multimorbidity, respectively. Compared with concordant multimorbidity, discordant multimorbidity was more prevalent among women, rural residents, former smokers, and individuals engaging in high-intensity physical activity. Discordant multimorbidity was also associated with poorer health outcomes and higher healthcare utilization, suggesting a greater burden in terms of management complexity and healthcare resource demand. These findings highlight the need for greater attention to the special needs of individuals with concordant and discordant multimorbidity in both research and clinical practice. Future studies with larger and more representative samples are warranted to further explore their potential implications for clinical decision-making.

## Data Availability

All data produced in the present study are available upon reasonable request to the authors

## Funding

This study was funded by the Research Unit of Evidence-Based Evaluation and Guidelines (2021RU017), Chinese Academy of Medical Sciences, School of Basic Medical Sciences, Lanzhou University, Lanzhou, Gansu, China.

### CRediT authorship contribution statement

Zjiun Wang: Conceptualization, Data curation, Formal analysis, Methodology, Visualization, Writing – original draft; Søren T. Skou: Writing – review and editing; Yaolong Chen: Funding acquisition, Supervision, Writing – review and editing; Janne Estill: Conceptualization, Methodology, Supervision, Writing – review and editing.

