## Appendix for "Multimorbidity Patterns and Associated Factors Among Middle-Aged and Older Adults in China: Evidence from the CHARLS Study"

Table A.1 Model Fit Indices for the Latent Class Analysis

| **Model** | **AIC** | **BIC** | **Log-likelihood** | **Entropy** |
| --- | --- | --- | --- | --- |
| 2-class | 114543.9 | 114749.9 | -57243.0 | 0.4491764 |
| 3-class | 113150.5 | 113463.0 | -56531.3 | 0.5610922 |
| 4-class | 112210.0 | 112629.0 | -56046.0 | 0.6313883 |
| **5-class** | **111813.2** | **112338.7** | **-55832.6** | **0.6670195** |
| 6-class | 111632.8 | 112264.9 | -55727.4 | 0.6525985 |
| 7-class | 111521.6 | 112260.2 | -55656.8 | 0.6774587 |
| 8-class | 111433.7 | 112278.9 | -55597.9 | 0.6843652 |

AIC, Akaike Information Criterion; BIC, Bayesian Information Criterion

Table A.2 Model Fit Indices for the Latent Class Analysis (only discordant multimorbidity)

| **Model** | **AIC** | **BIC** | **Log-likelihood** | **Entropy** |
| --- | --- | --- | --- | --- |
| 2-class | 92597.03 | 92797.03 | -46269.51 | 0.6781618 |
| 3-class | 91565.33 | 91868.77 | -45738.66 | 0.7299105 |
| **4-class** | **90673.26** | **91080.15** | **-45277.63** | **0.7467340** |
| 5-class | 90249.92 | 90760.26 | -45050.96 | 0.8068095 |
| 6-class | 89935.36 | 90549.14 | -44878.68 | 0.7833612 |
| 7-class | 89714.15 | 90431.38 | -44753.07 | 0.7116005 |
| 8-class | 89531.51 | 90352.19 | -44646.75 | 0.7404209 |

AIC, Akaike Information Criterion; BIC, Bayesian Information Criterion
